# Evaluation of the accuracy of a multi-infection screening test based on a multiplex immunoassay targeting imported diseases common in migrant populations

**DOI:** 10.1101/2023.07.24.23293073

**Authors:** Ruth Aguilar, Angeline Cruz, Alfons Jiménez, Alex Almuedo, Carme Roca Saumell, Marina Gigante Lopez, Oriol Gasch, Gemma Falcó, Ana Jiménez-Lozano, Angela Martínez-Perez, Consol Sanchez-Collado, Andrea Tedesco, Manuel Carlos López, María Jesús Pinazo, Thais Leonel, Zeno Bisoffi, Anna Färnert, Carlota Dobaño, Ana Requena-Méndez

## Abstract

**Background:** In this study we have evaluated the performance of a novel multiplex serological assay with a panel of 8 antigens able to simultaneously detect IgG to HIV, chronic hepatitis B (HBV) and C (HCV), Chagas disease, strongyloidiasis and schistosomiasis as a screening tool for imported diseases in migrants.

**Methods:** Six panels of 40 well-characterized, anonymized serum samples from individuals with the respective confirmed infections (n=240) were used as positive controls to assess the sensitivity of the multiplex assay. One panel of 40 sera from non-infected subjects were used to estimate the seropositivity cutoffs for each infection, and 32 additional non-infected sera were used as negative controls to estimate the sensitivity and specificity for each serology. The multi-infection screening test was validated in a prospective cohort of 48 migrants from endemic areas to assess assay performance.

The sensitivity of the Luminex assay was calculated as the proportion of positive test results over all positive samples by the primary reference test. The specificity was calculated using 32 negative samples. Uncertainty was quantified with 95% confidence intervals (CI) using receiver operating characteristic analyses.

**Results:** The sensitivity /specificity were 100%/100% for HIV (p41 antigen), 97.5%/100% (AUC:0.99,[95%CI: 0.96-1.00]) for HBV (core antigen), 100%/100% (AUC:1.00,[95%CI 1.00-1.00]) for HCV (core antigen), 92.5%/90.6%,(AUC:0.96,[95%CI 0.91-1.00]) for strongyloidiasis (31-kDa recombinant antigen (NIE)), 97.5%/100%,(AUC:0.97,[95%CI 0.93-1]) for schistosomiasis (combined serpin *Schistosoma mansoni* and *S.haematobium* antigens) and 92.5%/96.9%,(AUC: 0.96,[95%CI 0.92-1.00]) for Chagas disease ([*T.cruzi* kinetoplastid membrane protein-11 (KMP11)]).

In the migrant cohort, antibody response to KMP11 correctly identified 14/14(100%) individuals with Chagas disease, whereas HBV-core antigen and NIE-Strongyloides correctly identified 91.7% and 86.4% individuals with chronic hepatitis B and strongyloidiasis respectively.

**Conclusions:** We have developed a new 8-plex Luminex assay that is robust and accurate, and could facilitate the implementation of screening programmes for imported diseases in migrant populations.

## INTRODUCTION

Migration is a complex and growing global phenomenon, of critical importance to European, North American and other countries such as Australia and New Zealand (Western countries). Most migrants are disproportionately affected by certain infections such as tuberculosis, human immunodeficiency virus (HIV), hepatitis B virus (HBV) and hepatitis C virus (HCV), and multiple studies have reported higher prevalence of such infections among them compared with autochthonous populations.^1,2^ In 2020, 44% of new HIV diagnoses were attributed to migrants living in the European Union, particularly migrants from sub-Saharan Africa.^3^ Similarly, migrants from certain regions are at increased risk of hepatitis B and C whereas for other regions, the risk is even lower compared to the autochthonous population.^2^

Other parasitic infections, such as Chagas disease, strongyloidiasis and schistosomiasis, which are not endemic in Western countries, -with the exception of strongyloidiasis in few areas-, are highly prevalent among migrants^4–6^. Chagas disease, caused by *Trypanosoma cruzi* is a public health concern due to the high pooled prevalence (4.2%) among migrants from Latin-American endemic countries living in Europe^5^, which is particularly high in Bolivian migrants (18%)^7^; but also due to the possibility of autochthonous transmission (congenitally, by transfusion, or transplantation,^8^) with a subsequent increase of related morbidity in national health systems from Western countries, particularly in immunosuppressed patients due to the risk of reactivation.^9^ Strongyloidiasis is also an emerging infection which is of public health concern due to the possibility of being a life-threatening pathology when immunosuppression is established.^10^ Schistosomiasis is a neglected tropical disease becoming relevant in low- and non-endemic countries because of the increased migration flows from high-endemic countries. It represents a management challenge due to the lack of clinical awareness and knowledge among health professionals.^11^

Early detection and treatment of these infections can lower the related morbidity and mortality.^11^ The European Centre for Disease Prevention and Control (ECDC) guidance on screening of infections in migrants,^12^ recommends the screening of HIV, HBV, HCV, strongyloidiasis and schistosomiasis. Chagas disease was not included in the ECDC screening and vaccination recommendations document, yet this disease has an impact in Europe’s health system and some studies have demonstrated the cost-effectiveness of the strategy of screening migrants from endemic countries at primary care.^8^

The feasibility of innovative integrated screening programmes of infectious diseases, has been proven in different settings including primary care units in United Kingdom, United States, Canada and Spain.^11,13–15^

Antibody-based assays are used for assessing the exposure to different infections among migrants including HIV or viral hepatitis,^16^ and also parasites (*Strongyloides stercoralis*^17^*, Schistosoma* spp^18^ and *T. cruzi*^19^) for which methods aiming at the direct detection of the parasites or their DNA have lower sensitivity. Quantitative suspension array assays simultaneously detecting antibodies to multiple pathogens using a single specimen can support a more cost-effective implementation of integrated disease surveillance systems. In addition, rapid tests with single but also with multiple biomarkers are gaining popularity in low-income countries. In this regard, the dual rapid HIV and *Treponema pallidum* diagnostic test is an example of multiple-test that has been utilized in integrated screening systems.

Luminex technology is gaining traction in diagnosis because of the ease, high throughput, and minimal sample volume requirements. It has been used to perform the surveillance of tropical viral infections,^20^ enteropathogens,^21^ or malaria.^22^ More recently another new multiplex-test has been developed to detect HIV, viral hepatitis, herpes and *T. pallidum.*^23^

Our study aimed to develop a new multiplex serological assay using 10 antigens to quantify IgG to six pathogens that are prevalent in migrant populations living in Catalonia (Spain), including HIV, HBV, HCV, *T. cruzi, S. stercoralis* and *Schistosoma* spp., and to estimate its sensitivity and specificity compared with primary reference standard tests for each infection.

## METHODS

### Study design

The study was performed using six panels of archived, fully anonymized well-characterized coded serum samples previously diagnosed for the different infections, plus a group of samples from non-infected subjects.

Each panel consisted of 40 positive control specimens for each pathogen (HIV, HBV, HCV, *T. cruzi*, *Strongyloides* and *Schistosoma*). HIV positive controls were determined with the Determine HIV-1/2 Rapid Test (Abbott Laboratories) and confirmed with the Unigold rapid test (Trinity Biotech). HBV and HCV positive controls were determined by RNA quantitative testing that measures the viral load. The schistosomiasis positive controls were positive for stool and/or urine test depending on the species. The strongyloidiasis positive controls were individuals diagnosed with strongyloidiasis and included in a clinical trial that assessed the effectiveness of ivermectin^24^ and for which the inclusion criteria were having a positive faecal test for *S. stercoralis* or having a positive serological test at high titers irrespective of the result of faecal test. High titers were assessed as at least 160 titer for the indirect fluorescent antibody test (IFAT)-that is, whole serum dilution 1/160, and at least 2x normalized optical density(OD) for the IVD Research ELISA, -that it, the ration between OD of the sample and that of the weak positive control-^25^. The accuracy of these tests at these high titers have been validated showing that they approach 100% specificity, while maintaining 70% sensitivity.^25^ The *T. cruzi* positive controls were obtained from a serum biobank of Chagas cases defined by being positive in at least two serological tests using different recombinant antigens. For the parasitic infections, further details of the serological titers, and faecal or urine samples in case of strongyloidiasis and schistosomiasis are reported in Annex 1. From the panel of 72 sera of healthy subjects, 40 samples were used to estimate the seropositivity cutoffs for the different serologies, and 32 were used as negative controls to estimate the specificity for each serology. We validated the multi-infection screening test in a prospective cohort of 48 migrants with epidemiological exposure risk to the above infections depending on the country of origin. They were recruited during the implementation of a multi-disease screening program for asymptomatic migrants attended at primary care units in Catalonia, (January 2019 - December 2020). They were tested for each infection with the reference method depending on the exposure risk.

The reference serological tests used to assess the accuracy of the Luminex in the prospective migrant cohort were performed according to each centre’s referral laboratory for HIV and viral hepatitis; for *Strongyloides,* an enzyme-linked immuno-sorbent assay (ELISA) based on IVD-*S.stercoralis* crude antigen (SCIMEDX, Dover-NJ,) or an in-house immunofluorescence; for *Schistosoma,* an ELISA or an in-house indirect-haemagglutination test (Schistosomiasis-Fumouze, Levallois-Perret,); for *T. cruzi*, a commercial ELISA with recombinant antigens (BioELISA-Chagas, Biokit-SA), and an in-house ELISA with whole epimastigotes antigen^26^, with diagnosis defined by positivity in both serological tests.

### Multi-infection IgG Luminex assay

The 10 antigens initially selected for the study are listed in Table 1. P24 (core antigen) and gp41 (envelope antigen) were selected for HIV serology, the first one for being indicative of acute infection^27^ and the second one for being highly immunogenic during seroconversion^28^ the 31-kDa recombinant antigen (NIE) antigen was selected for the serology of *S. stercoralis*, as ELISAs based on this antigen have shown high sensitivity in the range of 71– 84%^25,29,30^ *S.mansoni* (Sm) and *S.haematobium* (Sh) serpins were selected for the serology of the corresponding species for being reported as promising species-specific diagnostic antigens by Luminex^31^, hepatitis C core and nonstructural protein 3 (NS3) antigens were tested for chronic hepatitis C infection, the core antigen is currently used for diagnostic^32^ and the NS3 antigen has also been reported as seroreactive^33^, hepatitis B core (anti-HBc) antigen was used for the detection of chronic hepatitis B infection as the presence of IgG indicates ongoing infection, and people who have immunity to hepatitis B from a vaccine do not develop anti-HBc^34^. Finally, paraflagellar rod proteins 2 (PFR2)1 and *T.cruzi* kinetoplastid membrane protein-11 (KMP11) antigens were selected for being previously reported as good markers of *T.cruzi* infection^35^.

**Table 1.** The 10 antigens tested used in the multi-infection Luminex assay.

| Antigen | Pathogen | Source | Coupling concentration | Reference |
| --- | --- | --- | --- | --- |
| Recombinant HIV1 p24 protein | HIV | Abcam ab43037 | 30µg/mL | <sup>37</sup> |
| Recombinant HIV1 gp41 protein | HIV | Abcam ab49068 | 250µg/mL | <sup>45</sup> |
| NIE | Strongyloides | Expressed in *E. coli* by Sukwan Handali (CDC) from a plasmid clone donated by Thomas Nutman (NIH/NIAID), USA | 10µg/mL | <sup>46</sup> |
| Sm serpin | *Schistosoma mansoni* | Donated by Satoshi Kaneko (Nagasaki University, Japan) | 30µg/mL | <sup>47</sup> |
| Sh serpin | *Schistosoma haematobium* | Donated by Satoshi Kaneko (Nagasaki University, Japan) | 50µg/mL | <sup>47</sup> |
| Recombinant Hepatitis C Virus NS3 protein | HCV | Abcam ab49024 | 50µg/mL | <sup>48</sup> |
| Recombinant Hepatitis C Virus Core Antigen | HCV | Abcam ab49017 | 50µg/mL | <sup>48</sup> |
| Recombinant Hepatitis B Virus Core Antigen | HBV | Abcam ab49013 | 30µg/mL | <sup>49, 50</sup> |
| Chagas PFR2 | *Trypanosoma cruzi* | Recombinant protein produced by Carmen Thomas and Manuel C. Lopez, IPBLN-CSIC, Granada, Spain | 50µg/mL | <sup>51</sup> |
| Chagas KMP11 | *Trypanosoma cruzi* | Recombinant protein produced by Carmen Thomas and Manuel C. Lopez IPBLN-CSIC, Granada, Spain) | 30µg/mL | <sup>51</sup> |
| HIV: Human Immunodeficiency Virus, HBV: Hepatitis B Virus, HCV: Hepatitis C Virus; Sm: *Schistosoma mansoni*; Sh: *Schistosoma haematobium*; NS3: non-structural protein 3; PFR2: paraflagellar rod protein 2; KMP11: kinetoplastid membrane protein-11 |  |  |  |  |

Each of the 10 antigens included in the Luminex panel was coupled to a specific magnetic microsphere region; the optimal bead-coupling concentration was determined for each using the same methodology, as described previously.^36^

A detailed description of the assays is provided in Annex 2. Briefly, 361 samples were tested together with serial dilutions of a positive control to generate a standard curve for assay quality control, plus 3 technical blanks. Antigen-coupled microspheres were added in multiplex to the 384-well plate (2000 beads/ antigen/ well) and mixed with test samples, positive controls, and blanks. Final dilution of test samples was 1/250. The plate was incubated for 1 h at room temperature in agitation and protected from light. Then, the plate was washed and 25μL of goat anti-human IgG-phycoerythrin,(1:400) were added to each well and incubated for 30 min. The plate was washed again and microspheres resuspended with Luminex Buffer, to be acquired on the Flexmap-3D® reader. At least 50 microspheres/analyte/well were acquired, and median fluorescence intensity (MFI) was reported. Annex 3 shows the standard curves made of 20 serial dilutions of the positive control. Reference standard results were blinded to the researchers who performed the Luminex test.

### Data analysis

MFI cutoffs for seropositivity were calculated for each antigen as the exponential function of the mean plus 2 standard deviations (SD) or 3SD of log_10_-transformed MFIs of 40 negative controls randomly selected among the specimens from 72 subjects with no previous exposure to any infection. The other 32 negative controls were used to estimate the specificity of the Luminex assay vs the reference tests. Thereafter, for each infection, the evaluation of the Luminex assay performance used 72 specimens with known serological status (40 positive and 32 negative samples). The sensitivity of the Luminex test was calculated as the proportion of positive results (based on the cut-off with 2SD) over all positive samples by the primary reference test. Sensitivity was also calculated with the cutoff with 3SD. Similarly, specificity was calculated over 32 control negative samples from the group of healthy controls using both 2SD and 3SD cutoffs. Uncertainty was quantified with 95% confidence intervals (CI) using receiver operating characteristic (ROC) analyses. Finally, the sensitivity and specificity were also estimated in a panel of samples prospectively collected from 48 migrants at risk of infections.

### Ethics

Positive and negative control samples used to develop the assay were obtained from collections registered at the Instituto de Salud Carlos III, Hospital Clínic(Barcelona)/ISGlobal biobanks and Sacro-Cuore Hospital Tropica-Biobank (Annex 4). The study protocol and informed consent from migrants whose samples were prospectively collected were approved by the ethical committee at Hospital Clínic, (HCB/2017/0847). Results are reported according to the STARD-checklist (Annex 5).

## RESULTS

### Accuracy of the multi-infection Luminex assay on a reference panel of human samples

We evaluated the performance of the novel multiplex serological assay on a panel of human sera with known infection status. First, we estimated the seropositivity cutoffs for each of the antigens in the Luminex panel using 2 and 3SD, summarized in Annex 6.

Figure 1 represents the IgG serological levels (log_10_MFI) against each antigen in the Luminex panel in 32 negative and 246 positive control samples (1A), and 48 test samples from migrants exposed to the infections (1B).

**Figure 1.**
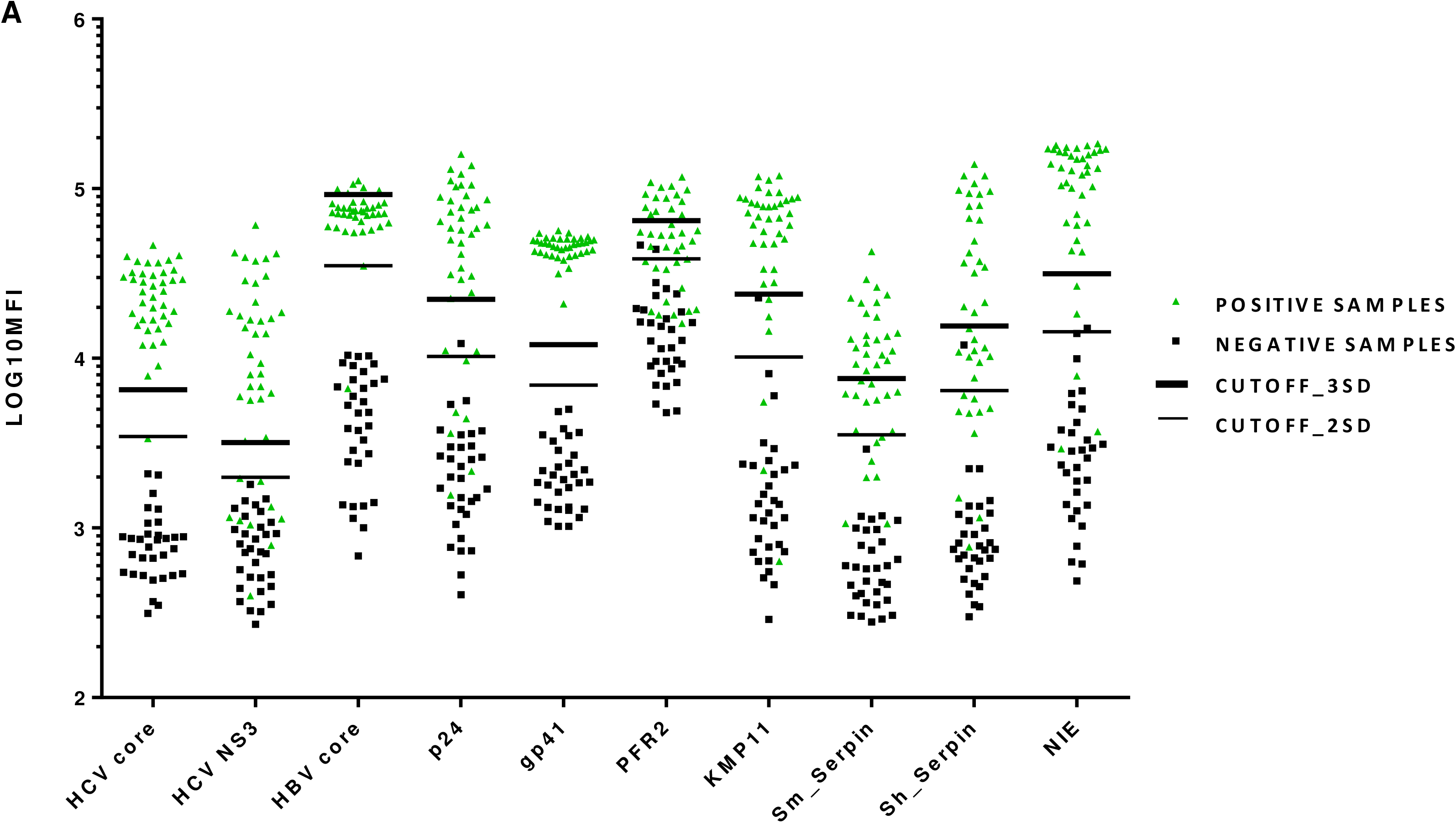
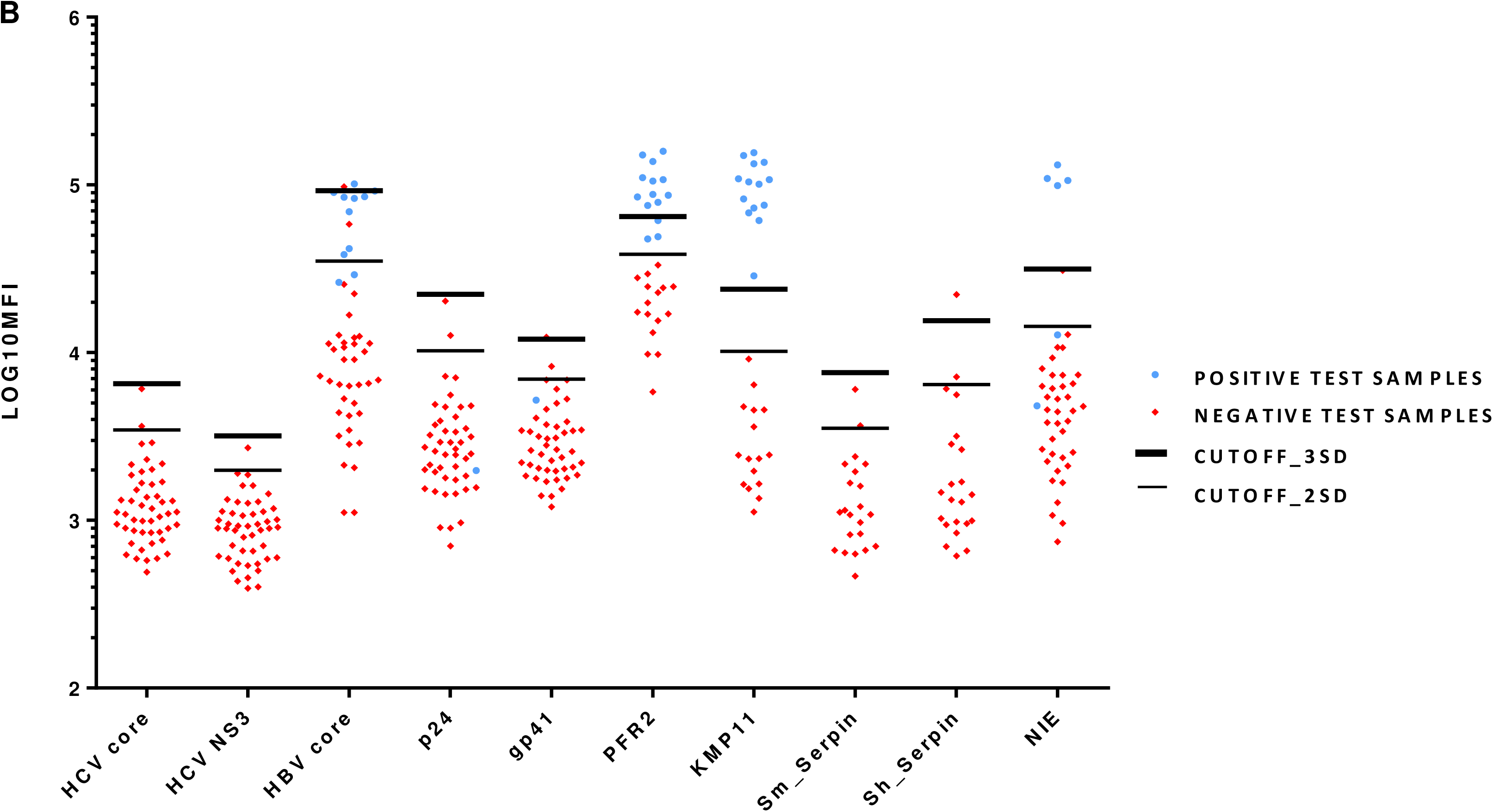
Dot plots showing the distribution of IgG levels measured against the 10 multiplexed antigens in the different panels of samples. 1A. Panels of positive and negative control samples for each infection 1B. Panel of test samples from the migrants cohort Legend: The positive samples (1A) were selected based on seropositivity in the reference methods. In total, 40 positive samples were included for the respective infections (n=240). Log10MFI: IgG serological levels (log10 mean fluorescence intensity); HBV: Hepatitis B Virus, HCV: Hepatitis C Virus: Sm: *Schistosoma mansoni*; Sh: *Schistosoma haematobium*

Using a 2SD cutoff, the sensitivity was generally high (>90%) for the majority of the antigens, whereas the specificity depended on the pathogen and antigen (Table 2 and Figure 2). The sensitivity and specificity of the Luminex test for detecting HIV using the gp41 recombinant protein determined by ROC analysis were 100% using both 2SD and 3SD cutoffs. In contrast, the p24 protein performed worse, with a sensitivity of 87.5% and a specificity of 96.9% with the 2SD cutoff, and 80% sensitivity at 3SD cutoff (Figure 2A).

**Figure 2.**
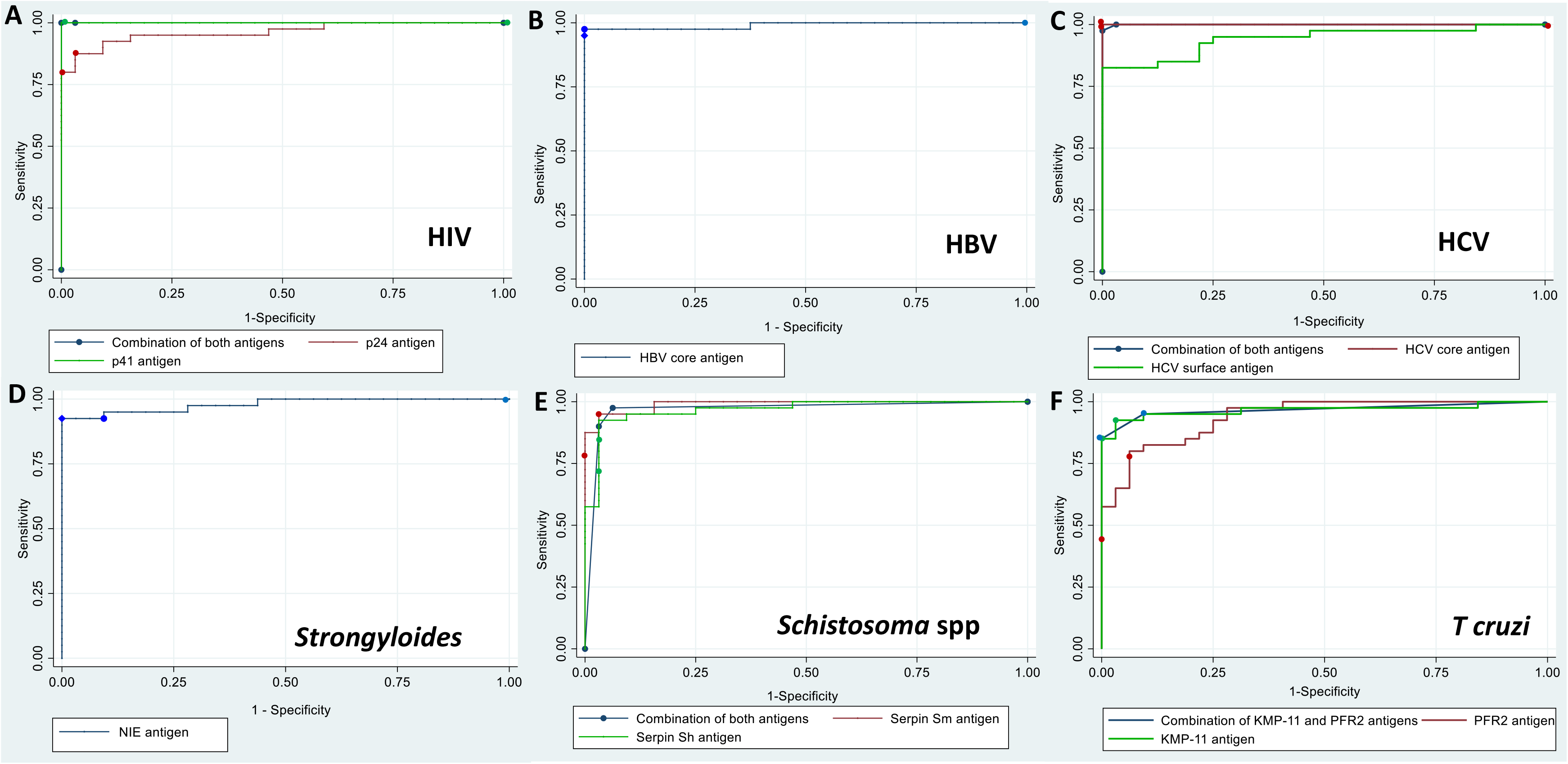
Receiver operating curves for the 10 antigens tested in the multiplex assay to detect the infections.

**Table 2.**
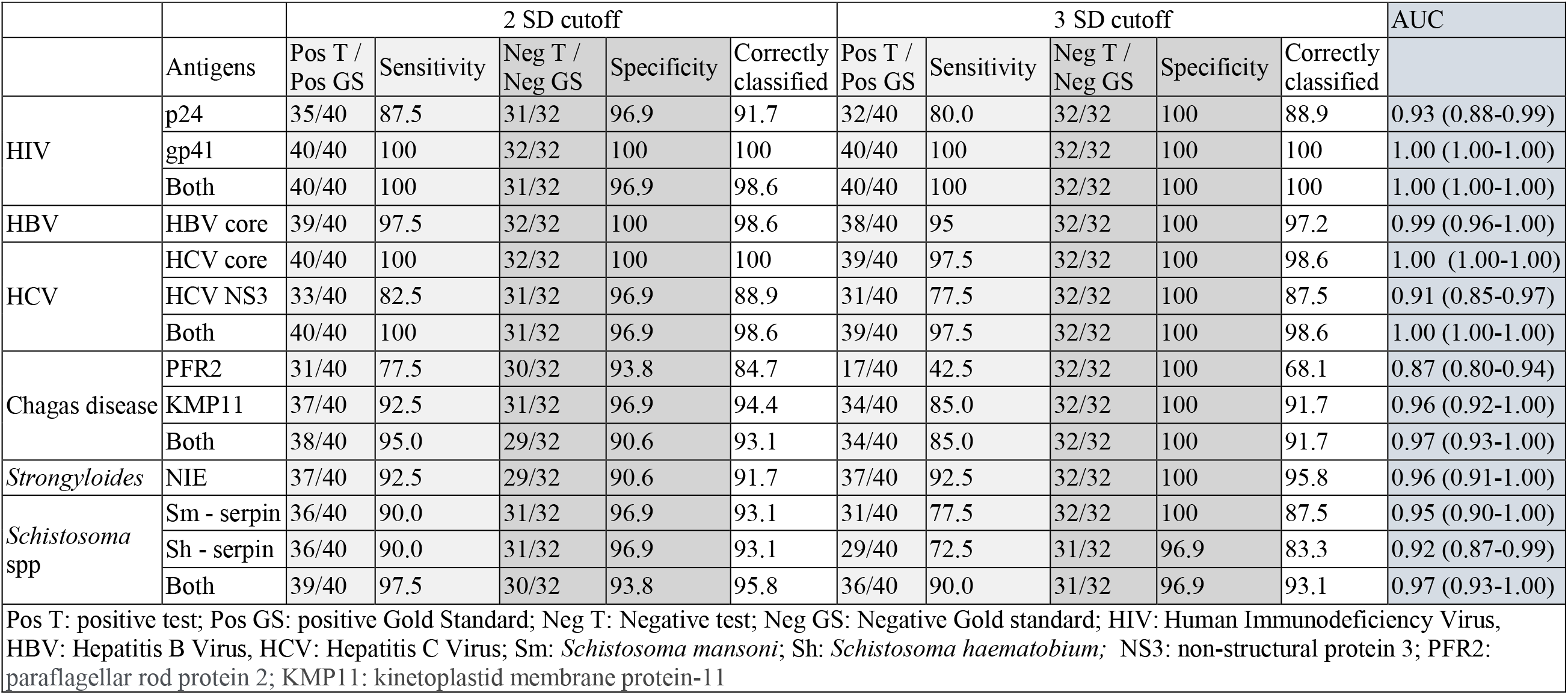
Performance parameters of the 10 antigens tested in the multiplex assay on a reference panel of human samples.

The HBV core antigen showed a sensitivity of 97.5% at 2SD-cutoff and 95% at 3SD-cutoff, and a specificity of 100% (2SD and 3SD-cutoff), (AUC: 0.99, [95%CI: 0.96-1.00]), (Figure 2B). The HCV core antigen showed a 100% sensitivity at 2SD-cutoff (97.5% at 3SD-cutoff) and a specificity of 100% (2SD and 3SD-cutoff),(AUC: 1.00,[95%CI 1.00-1.00]), (Figure 2C).

*Strongyloides*-NIE antigen reported a sensitivity of 92.5% (2SD and 3SD-cutoff) and specificities of 90.6% (2SD-cuttoff) and 100% (3SD-cutoffs), (AUC:0.96, [95%CI 0.91-1.00]), (Figure 2D). In a restricted analysis (Annex 7) to those samples that tested positive in feces [(either by polymerase chain reaction (PCR) test or any other direct technique]), the performance of the NIE antigen in Luminex with the 3SD cutoff improved to 100% sensitivity (AUC: 1.00, [95%CI 1.00-1.00]).

The sensitivity and specificity of the Luminex test to detect schistosomiasis was 97.5% and 93.8% (2SD cutoff),(AUC:0.97, [95%CI 0.93-1.00]) using *S. haematobium* (Sh) *and S. mansoni* (Sm) serpin antigens together, (Figure 2E). Interestingly, the sensitivity of the serpin-Sh antigen was 100% for detecting *S. haematobium* infections whereas the sensitivity of the serpin-Sm antigen was 96% for detecting *S. mansoni* infections (Annex 7).

The *T. cruzi* KMP11 antigen performed better (Sen 92.5%, Spec 96.9%, AUC:0.96, [95%CI 0.92-1.00]) compared to the PFR2 antigen (Sen 77.5%; Spec 93.8, AUC:0.87, [95%CI 0.80-0.94]). Furthermore, when combining both antigens, the sensitivity increased to 95% (AUC:0.97, [95%CI 0.93-1.00]) (Figure 2F). There were two samples in the *T. cruzi* infection panel that resulted indeterminate by the reference test, and it was necessary a third serological test to confirm the infection. After excluding these samples from the analysis (Annex 7), the sensitivity increased to 97.4% at 2SD cutoff (AUC:0.98, [95%CI 09.95-1.00]).

In order to assess possible cross-reactivities, we evaluated for each infection the false positive rate among the individuals from the six panels of positive controls (Annex 8). Overall, the Luminex serology showed a low frequency of false positives, except for *S. stercoralis* showing a rate of 66% (12/18) and 50% (9/18) among *Schistosoma* spp. Infections at a cutoff of 2SD and 3SD respectively, and 62% (13/21) and 38% (8/21) in *Schistosoma* spp. and HBV coinfections at a cutoff of 2SD and 3SD respectively. There were 25% (5/20) false positives for HIV and 36% (4/11) for HCV in *Schistosoma* spp. and HBV coinfections at a cutoff of 2SD that were not observed at 3SD.

### Accuracy of the multi-infection Luminex assay on migrant’s cohort

There were no positive reference tests for HIV, HCV, and schistosomiasis among the migrant’s cohort, therefore the sensitivity and AUC of the Luminex test for these infections could not be evaluated. The specificities assessed for each of these infections are detailed in table 3.

**Table 3.** Performance parameters of the 10 antigens tested in the multiplex assay in a prospective cohort of 48 migrant individuals with risk of exposure to the tested infections.

|  |  | 2 SD cutoff |  |  |  |  | 3 SD cutoff |  |  |  |  | AUC |
| --- | --- | --- | --- | --- | --- | --- | --- | --- | --- | --- | --- | --- |
|  | Antigens | Pos T /<br>Pos GS | Sensitivity | Neg T /<br>Neg GS | Specificity | Correctly<br>classified | Pos T /<br>Pos GS | Sensitivity | Neg T /<br>Neg GS | Specificity | Correctly<br>classified |  |
| HIV | p24 | 0/1 | 0 | 43/45 | 95.6 | 93.5 | 0/1 | 0 | 43/45 | 95.6 | 93.5 | 0.48 ( . - 1.00) |
|  | gp41 | 1/1 | 100 | 39/45 | 86.7 | 87.0 | 0/1 | 0 | 44/45 | 97.8 | 95.7 | 0.92 ( . - 1.00) |
| HBV | HBV core | 11/11 | 100 | 33/37 | 89.2 | 91.7 | 7/11 | 63.6 | 35/37 | 94.6 | 87.5 | 0.95 (0.90-1.00) |
| HCV | HCV core | - | - | 46/48 | 95.8 | - | - | - | 47/48 | 97.9 | - | - |
|  | HCV NS3 | - | - | 45/48 | 93.8 | - | - | - | 45/48 | 93.8 | - | - |
| Chagas disease | PFR2 | 14/14 | 100 | 16/16 | 100 | 100 | 12/14 | 85.7 | 16/16 | 100 | 93.3 | 1.00 (1.00-1.00) |
|  | KMP11 | 14/14 | 100 | 16/16 | 100 | 100 | 14/14 | 100 | 16/16 | 100 | 100 | 1.00 (1.00-1.00) |
|  | Both | 14/14 | 100 | 16/16 | 100 | 100 | 14/14 | 100 | 16/16 | 100 | 100 | 1.00 (1.00-1.00) |
| *Strongyloides* | NIE | 5/6 | 83.3 | 33/38 | 86.8 | 86.4 | 4/6 | 66.7 | 37/38 | 97.4 | 93.2 | 0.88 (0.70-1.00) |
| *Schistosoma*<br>spp | Sm - serpin | - | - | 20/22 | 90.0 | - | - | - | 21/22 | 95.4 | - | - |
|  | Sh - serpin | - | - | 18/22 | 81.8 | - | - | - | 21/22 | 95.4 | - | - |
|  | Both | - | - | 18/22 | 81.8 | - | - | - | 21/22 | 95.4 | - | - |

Pos T: Positive test; Pos GS: Positive Gold Standard; Neg T: Negative test; Neg GS: Negative Gold standard; HIV: Human Immunodeficiency Virus, HBV: Hepatitis B Virus, HCV: Hepatitis C Virus; Sm: *Schistosoma mansoni*; Sh: *Schistosoma haematobium*; NS3: non-structural protein 3; PFR2: paraflagellar rod protein 2; KMP11: kinetoplastid membrane protein-11

Among the 48 individuals tested for HBV, 11 of them had a positive anti-HBc test result. The core HBV antigen tested in Luminex showed a sensitivity of 100% and specificity of 89.2% (2SD-cutoff). For the KMP11-Chagas antigen, the Luminex assay correctly identified (2SD and 3SD-cutoffs) the 14 positive and 16 negative cases detected with the reference method (100% Sens and 100% Spec, AUC: 1.00, [95%CI 1.00-1.00]). The NIE-antigen showed 83.3% sensitivity and 86.8% specificity, at 2SD-cutoff.

We also assessed for cross-reactivities among the 48 individuals from the migrant’s cohort (Annex 9), and frequency of false positives was overall low, except for *S. stercoralis infections* with 20% (2/10) false positives among *T. cruzi* but only at 2SD and 28% (2/7) among HBV infections at 2SD and 15% (1/7) at 3SD cutoff. HIV showed 32% (4/12) false positives among the *T. cruzi* infected individuals at 2SD (only 1 at 3 SD), and *Schistosoma* spp. showed 28% (2/7) among HBV infected individuals, using the 2SD and 2 SD cutoff.

## DISCUSSION

We have developed a 8-plex Luminex assay for the simultaneous detection of IgG against HIV, HBV, HCV, *Schistosoma* spp, *S. stercoralis* and *T. cruzi*. To our knowledge, this is the first multiplex test that simultaneously detects several viral and parasitic infections based on serological tests that are prevalent in migrant populations. Although 10 antigens were initially evaluated, only 8 of them showed a good performance.

Our 8-plex assay results showed a good agreement with the reference tests, being highly accurate to detect most pathogens, particularly gp41-HIV and HCV-core antigen, (100% sensitivity), the combination of *Schistosoma* serpin antigens (97.5%), HBV core antigen (97.5%) and the combination of *T. cruzi* antigens (95%).

For the infections for which two antigens were tested, the sensitivity of the test increased for Chagas disease and schistosomiasis, whereas for HIV the combination of p24 and gp41 antigens slightly decreased the specificity of the test compared with the use of the gp41 antigen alone. P24 antigen tends to negativize over time after the infection^37^, as it is only detected in blood during early stages of the primoinfection, thus it is not reliable for screening except for the detection of recent infections.^38^ Similarly to other studies,^39^ for HCV the combination of NS3 with the core antigen decreased the specificity of the test compared with the core antigen alone. The performance of HIV and HBV antigens was similar compared with a previous Luminex study, whereas the core HCV antigen has demonstrated better performance.^23^ However HIV gp41 and HCV-core antigens showed some false positives in *Schistosoma* spp. and HBV coinfections, and HIV also in *T.cruzi* infections, suggesting some room for improvement.

As previously described, the performance of the recombinant *Strongyloides*-NIE antigen (sensitivity 92.5%) is not as good as other commercially available tests based on crude antigens.^25^ However, the sensitivity was 100% for those infections detected by microscopy or PCR. Therefore, the lower sensitivity of NIE by Luminex can be attributed to a poorer diagnostic capability of the recombinant antigen compared with crude antigens.^7^ However, the *Strongyloides* serology with crude antigen may give false positives due to cross-reactivity with other helminths, such as, filarial infections, *Ascaris lumbricoides* infection, and acute schistosome infections^40^, thus these specimens could also be true negatives wrongly characterized by the reference test. In fact, when assessing the rate of false positives due to possible cross reactivities, *S. stercoralis* showed a high prevalence of false positives among other infections suggesting the need to improve the specificity for the NIE antigen.

Concerning Chagas disease, although the combination of both antigens showed a better sensitivity, the KMP11 antigen performed much better compared with PFR2 antigen to detect *T. cruzi*, improving even the accuracy when excluding borderline results detected by the reference test, which suggests that this antigen is performing similar compared with other serological tests developed for *T. cruzi* screening.^41^ In any case, the diagnosis of Chagas disease by Luminex should be confirmed with another serological test using a different recombinant protein, as standard procedure.

Interestingly, when the Luminex assay was tested in a prospective sample of migrant individuals coming from countries endemic for the infections tested, the assay showed a good accuracy to detect most infections, although sensitivity was lower for strongyloidiasis, suggesting that this Luminex panel may be promising as a screening tool. However, the migrants sample size was very small and there were no patients with patients with schistosomiasis, HIV, and hepatitis C.

We have tested the performance of the 10 antigens using two different cutoffs. The 2SD-cutoff gives higher sensitivities by losing some specificity, while the 3SD-cutoff gives lower sensitivities but 100% specificity for all infections. In the context of screening of migrant populations, higher sensitivities are more important than 100% specificities, as having infected undiagnosed individuals is not desirable, therefore false negative cases should be minimized.

Overall, the use of bead-based multiplex assays is gaining recognition in diagnosis of infections because of the ease, high throughput, and minimal sample/reagent volume requirements^42–44^ allowing measuring up to 100 analytes/per sample, and the FlexMap-3D platform (the one used in this study) up to 500 analytes per sample, with comprehensive reductions in time and cost. Remarkably, multiplexing more than one antigen may increase the sensitivity of pathogen detection compared to single-biomarkers. In addition, Luminex panels are flexible and can be adapted to the epidemiological risk of individuals (i.e., the country of origin). Thus, for example *T cruzi* antigen could be only included in a specific Luminex panel for Latin American migrants.

In this specific study, we evaluated the diagnostic utility of a multiplex test focusing on the positivity or negativity of samples, however antibody levels could be useful in studies addressing IgG kinetics since time of infection, or IgG levels depending on viral or parasitic loads.

Although the Luminex technology itself is not novel, to our knowledge, this is the first multiplex test that simultaneously detects several viral and parasitic infections that are prevalent in migrant populations based on serological tests. The simultaneous classification for the presence or absence of multiple pathogens in a single assay has the potential to revolutionize and simplify the public health and diseases surveillance in endemic areas where the infections are prevalent,^44^ and also the implementation of screening programmes in migrant populations. Moreover, 5 of the 10 proteins used in the panel are not commercially available thus have been produced in house, and for some of them this is the first time that their diagnostic performance is reported as part of a multi-diagnostic infection test.

In addition, the 8-plex assay overcomes many of the laboratory infrastructure requirements of the single infection diagnostic testing, thereby simplifying the logistics. This makes it a more accessible test for settings that may not have either the technical knowledge or the infrastructure capacity to conduct the single diagnostic tests using a dedicated platform for each of them (i.e., primary care). In this regard, the current existence of benchtop compact easy-to-use multiplex equipment, would facilitate the development of a point of care version of the assay in primary care.

While further work is needed to optimize and validate this 8-plex assay, and maybe expand it to other infections, and to other assays using non-invasive samples such as mucosa or saliva, it has the potential to drastically reduce the disease burden in settings where testing has often been limited, particularly for imported parasitic diseases.

A detailed comparative cost-analysis between the multiplex assay and the individual standard conventional tests for each infection is in progress, this test has substantial potential to improve diagnostic testing, based on this proof-of-concept study.

The main limitation of the study is the small sample size of the prospective sample with no diagnosed cases for some of the infections. For this reason, we plan to conduct a more robust prospective study for the validation of the 8-plex assay. Another limitation of the study is the lack of a proper challenge panel made up of samples from infections and conditions that may cause potential cross-reactivity (i.e., filariasis in the case of strongyloidiasis)^7^. The reason was the lack of samples with known unique infections for this purpose. Instead, we have used the data from the individuals from the six panels of positive controls and the 48 tested individuals with information on other infections as challenge panels. We are measuring IgG, which is indicative of exposure, not distinguishing between current or past infection.

However, the Luminex platform allows measuring any isotype or IgG subclass, thus IgM or IgA could be used in combination with IgG to distinguish recent from past infections. In addition, the use of specific antigens can be helpful to determine the approximate time of infection.

## Conclusions

We present a proof-of-principle of the excellent performance of a high through put single test using multiplexed antigens with a minimal sample volume that can detect multiple infections, common in migrant populations. Our 8-plex assay has shown to be robust and accurate, although further prospective study should validate the reproducibility of the assay. This test could facilitate the implementation of screening programmes particularly in non-hospital-based settings where the detection of single pathogens can be logistically challenging and expensive.

## Data Availability

All data produced in the present study are available upon reasonable request to the authors

## Acknowledgements

We are grateful to all patients who participated in this study, the general practitioners, nurses, and other staff from the primary care centres and hospitals who were committed to perform the field work required for the study while maintaining their daily tasks. Specially, we acknowledge the work carried by Maria Luisa Machado and Silvia Capilla from Hospital Universitari Parc Taulí. Special thanks to the research group at the IRCCS Sacro Cuore Don Calabria Hospital for providing the positive control samples for strongyloidiasis (part of them) and schistosomiasis, in particular to Stefano Tais and Monica Degani, to Jose Munoz (Hospital Clinic, Barcelona) for facilitating positive control samples for strongyloidiasis (part of them) and Denise Naniche (ISGlobal) for information on the HIV positive control samples. We are also grateful to Mª Carmen Thomas from the Institute of Parasitology and Biomedicine López Neyra for supplying the Chagas disease antigens, Thomas Nutman (NIH/NIAID,USA), Sukwan Handali (CDC) for supplying the NIE, Satoshi Kaneko (Nagasaki University, Japan) for donating the Sm and Sh serpins, and Marta Vidal, Rebeca Santano and Inocencia Cuamba for support in the antigen procurement and characterization.

## Author contribution

Study design by ARM, RA, AJ and CD. Field work done by CRS, MGL, OGB, GF, AJL, AMP, RSC. Sample processing carried out by AJ, RA and CD. Methodology developed by RA, AC and ARM. Interpretation of results executed by AC, ARM and AJ. The manuscript has been drafted by RA, AC, AJ, CD and ARM. All authors have contributed to the writing, reviewing and editing of the publication.

## Grants that funded the project

This study was supported by two grants from Instituto de Salud Carlos III (ISCIII), through the “Fondo de Investigación para la Salud (FIS)”, PI17/02020 and PI20/00866.

ARM receives funding from the Strategic Research Program in Epidemiology at Karolinska Institutet.

The research team was supported by CIBER-Consorcio Centro de Investigación Biomédica en Red-(CB 2021).

MCL was supported by the Programa Estatal I+D+I, Spanish Ministry of Science and Innovation and FEDER (PID2019-109090RB-I00/AEI/10.13039/501100011033).

Authors from IRCCS Sacro Cuore Don Calabria Hospital were supported by the Italian Ministry of Health “Fondi Ricerca corrente-L2P4”.

Part of the data from this study were presented in the European Conference of Clinical Microbiology and Infectious Diseases in Copenhagen (April 2023).

