## supplementary material for "Evaluation of the accuracy of a multi-infection screening test based on a multiplex immunoassay targeting imported diseases common in migrant populations"

### List of Annexes

Annex 1. Test results of samples used in the positive control panel.

Annex 2. The multi-infection IgG Luminex assay

Annex 3. Standard curve generated with 20 serial dilutions (1:2) of a positive control for quality control

Annex 4. List of biobanks or collections from which the control samples of the assay have been obtained

Annex 5. STARD checklist (Standards for Reporting Diagnostic accuracy studies)

Annex 6. Seropositivity cutoffs for each of the antigens in the Luminex panel using 2 and 3 SD

Annex 7. Sensitivity and AUC analyses including only confirmed cases with parasitological techniques for each infection as positive controls.

Annex 8. Challenge panel 1: False positives of the Luminex assay in a panel of human samples tested positive for other infections.

Annex 9. Challenge panel 2: False positives of the Luminex assay in a prospective cohort of 48 migrant individuals tested positive for other infections.

**Annex 1. Test results of the samples used in the positive control panel.**

| *Trypanosoma cruzi* | | | |
|---|---|---|---|
| | **Serology 1** | **Serology 2** | **Sample year** |
| TS_CHA01 | 10,9 | 28,49 | 2017 |
| TS_CHA02 | 9,83 | 26,69 | 2017 |
| TS_CHA03 | 3,17 | 9,81 (doubtful) | 2017 |
| TS_CHA04 | 9,38 | 26,7 | 2017 |
| TS_CHA05 | 11,04 | 42,68 | 2017 |
| TS_CHA06 | 8 | 20,8 | 2016 |
| TS_CHA07 | 11,37 | 36,49 | 2017 |
| TS_CHA08 | 9,88 | 25,49 | 2016 |
| TS_CHA09 | 12,14 | 39,76 | 2017 |
| TS_CHA10 | 11,78 | 35,84 | 2016 |
| TS_CHA11 | 7,6 | 22,19 | 2017 |
| TS_CHA12 | 10,45 | 25,22 | 2017 |
| TS_CHA13 | 8,95 | 20,31 | 2017 |
| TS_CHA14 | 11,32 | 23,64 | 2017 |
| TS_CHA15 | 2,01 | 7,2 (doubtful) | 2016 |
| TS_CHA16 | 12,79 | 30,87 | 2016 |
| TS_CHA17 | 1,05 | 12 | 2016 |
| TS_CHA18 | 2,61 | 11,84 | 2017 |
| TS_CHA19 | 10,67 | 28,29 | 2022 |
| TS_CHA20 | 12,3 | 42,14 | 2017 |
| TS_CHA21 | 14,21 | 26,23 | 2018 |
| TS_CHA22 | 10,96 | 16,67 | 2018 |
| TS_CHA23 | 6,94 | 27,22 | 2017 |
| TS_CHA24 | 11,7 | 32,22 | 2017 |
| TS_CHA25 | 13,81 | 27,15 | 2018 |
| TS_CHA26 | 12,06 | 20,4 | 2016 |
| TS_CHA27 | 11,32 | 11,6 | 2018 |
| TS_CHA28 | 10,65 | 23,64 | 2018 |
| TS_CHA29 | 10,24 | 18,31 | 2018 |
| TS_CHA30 | 7,68 | 23 | 2016 |
| TS_CHA31 | 11,61 | 23,4 | 2018 |
| TS_CHA32 | 10,32 | 21,97 | 2017 |
| TS_CHA33 | 6,8 | 20,19 | 2018 |
| TS_CHA34 | 9,63 | 27,18 | 2017 |
| TS_CHA35 | 11,83 | 8,9 | 2018 |
| TS_CHA36 | 9,83 | 20,3 | 2018 |
| TS_CHA37 | 10,89 | 14,78 | 2019 |
| TS_CHA38 | 11,8 | 19,66 | 2019 |
| TS_CHA39 | 8,72 | 18,9 | 2017 |
| TS_CHA40 | 12,08 | 21,01 | 2017 |

| *Strongyloides stercoralis* | | | | | |
|---|---|---|---|---|---|
| | **Serology ELISA** | **Immunofluorescence** | **Faecal test** | **PCR** | **Date** |
| TS_STR01 | 10,195 | | Negative | | 2014 |
| TS_STR02 | 7,39 | | Positive | | 2014 |
| TS_STR03 | >14 | | Negative | | 2014 |
| TS_STR04 | 3,48 | | Negative | | 2014 |
| TS_STR05 | 4,135 | | Negative | | 2014 |
| TS_STR06 | 7,01 | | Positive | | 2014 |
| TS_STR07 | 1,8 | | Positive | | 2014 |
| TS_STR08 | 1,82 | | Positive | | 2014 |
| TS_STR09 | 4 | | Negative | | 2015 |
| TS_STR10 | 5,62 | | Negative | | 2015 |
| TS_STR11 | 4,41 | | Positive | | 2015 |
| TS_STR12 | 3,29 | | Negative | | 2015 |
| TS_STR13 | 13,75 | | Negative | | 2015 |
| TS_STR14 | 4 | | Positive | | 2015 |
| TS_STR15 | 14 | | Positive | | 2015 |
| TS_STR16 | 7,28 | | Negative | | 2015 |
| TS_STR17 | 11,95 | | Negative | | 2015 |
| TS_STR18 | 9,01 | | Positive | | 2015 |
| TS_STR19 | 12 | | Negative | | 2015 |
| TS_STR20 | 6,43 | | Negative | | 2015 |
| TS_STR21 | 3 | | Negative | | 2015 |
| TS_STR22 | 12,3 | | Negative | | 2016 |
| TS_STR23 | 11,68 | | Negative | | 2016 |
| TS_STR24 | 6,79 | | Negative | | 2016 |
| TS_STR25 | 8,23 | | Negative | | 2016 |
| TS_STR26 | 9,9 | | Negative | | 2016 |
| TS_STR27 | 14 | | Positive | | 2016 |
| TS_STR28 | 4,86 | | Negative | | 2016 |
| TS_STR29 | 7 | | Positive | | 2016 |
| TS_STR30 | 13,55 | | Positive | | 2016 |
| TS_STR31 | 12,3 | | Negative | | 2016 |
| TS_STR32 | 3,22 | | negative | | 2016 |
| TS_STR33 | | 640 | | Positive | 2016 |
| TS_STR34 | 2,44 | 160 | Positive | Positive | 2016 |
| TS_STR35 | | 320 | Positive | | 2016 |
| TS_STR36 | | 80 | | Positive | 2016 |
| TS_STR37 | 3,24 | 160 | | Positive | 2016 |
| TS_STR38 | 1,53 | 160 | Positive | | 2016 |
| TS_STR39 | 2,67 | 160 | Positive | Positive | 2016 |
| TS_STR40 | | 640 | | Positive | 2016 |

| ***Schistosoma* spp** | | | | | | |
|---|---|---|---|---|---|---|
| | **Serology ELISA** | **CCA** | **Urine test** | **Faecal test** | **PCR** | **Date** |
| TS_SCH01 | 1,93 | Positive | Negative | *S. mansoni* | *Schistosoma* spp | 2016 |
| TS_SCH02 | 1,35 | Negative | Negative | *S. mansoni* | Negative | 2016 |
| TS_SCH03 | 1,815 | Positive | Negative | *S. mansoni* | *Schistosoma* spp | 2016 |
| TS_SCH04 | 2,5 | Negative | Negative | *S. mansoni* | Negative | 2016 |
| TS_SCH05 | 1,12 | Negative | Negative | *S. mansoni* | *Schistosoma* spp | 2016 |
| TS_SCH06 | 1,47 | Positive | Negative | *S. mansoni* | | 2016 |
| TS_SCH07 | 2,8 | Positive | Negative | *S. mansoni* | *Schistosoma* spp | 2016 |
| TS_SCH08 | 1,95 | Positive | Negative | *S. mansoni* | *Schistosoma* spp | 2016 |
| TS_SCH09 | 1,16 | Positive | Negative | *S. mansoni* | *Schistosoma* spp | 2016 |
| TS_SCH10 | 1,66 | Positive | Negative | *S. mansoni* | Negative | 2016 |
| TS_SCH11 | 1,64 | Positive | Negative | *S. mansoni - G. intestinalis* | Negative | 2016 |
| TS_SCH12 | 2,26 | Positive | Negative | *S. mansoni* | *Schistosoma* spp | 2016 |
| TS_SCH13 | 2,02 | Positive | Negative | *S. mansoni* | Negative | 2016 |
| TS_SCH14 | 3,74 | Positive | Negative | *S. mansoni* | *Schistosoma* spp | 2016 |
| TS_SCH15 | 1,765 | Positive | Negative | *S. mansoni* | Negative | 2016 |
| TS_SCH16 | 2,23 | Positive | Negative | *S. mansoni* | *Schistosoma* spp | 2016 |
| TS_SCH17 | 2,44 | Positive | Negative | *S. mansoni* | *Schistosoma* spp | 2016 |
| TS_SCH18 | 0,96 | Positive | Negative | *S. mansoni* | Negative | 2016 |
| TS_SCH19 | 1,66 | Positive | Negative | *S. mansoni* | *Schistosoma* spp | 2016 |
| TS_SCH20 | 3,37 | Positive | Negative | *S. mansoni* | *Schistosoma* spp | 2016 |
| TS_SCH21 | 1,77 | Doubtful | *S. haematobium* | Negative | Negative | 2016 |
| TS_SCH22 | 2,39 | Positive | *S. haematobium* | Negative | | 2016 |
| TS_SCH23 | 3,45 | Doubtful | *S. haematobium* | Negative | Negative | 2016 |
| TS_SCH24 | 1,81 | Negative | *S. haematobium* | | Negative | 2016 |
| TS_SCH25 | 1,1 | Negative | *S. haematobium* | *G. intestinalis* | *G.intestinalis* | 2016 |
| TS_SCH26 | 2,46 | Negative | *S. haematobium* | Negative | | 2016 |
| TS_SCH27 | 2,77 | Positive | *S. haematobium* | *S. mansoni* | *Schistosoma* spp | 2016 |
| TS_SCH28 | 0,88 | Positive | *S. haematobium* | Negative | Negative | 2016 |
| TS_SCH29 | 1,16 | Negative | *S. haematobium* | Negative | | 2016 |
| TS_SCH30 | 2,25 | Positive | *S. haematobium* | *S. hematobium* | | 2016 |
| TS_SCH31 | 2,88 | | | *S. mansoni* | | 2018 |
| TS_SCH32 | 3,63 | | *S. haematobium* | | | 2018 |
| TS_SCH33 | 1,87 | | *S. haematobium* | | | 2018 |
| TS_SCH34 | 3,29 | | | *S. mansoni* | | 2017 |
| TS_SCH35 | 1,6 | | *S. haematobium* | | | 2017 |
| TS_SCH36 | 2,74 | | *S. haematobium* | | | 2017 |
| TS_SCH38 | 2,37 | | | *S. mansoni* | | 2017 |
| TS_SCH37 | 3,89 | | *S. haematobium* | *S. mansoni* | | 2017 |
| TS_SCH39 | 1,92 | | | *S. mansoni* | | 2017 |
| TS_SCH40 | 2,63 | | | *S. mansoni* | | 2017 |

**Annex 2. The multi-infection IgG Luminex assay**

Each of the 10 antigens included in the Luminex panel (Table 1) was coupled to a specific magnetic microsphere region; the optimal bead-coupling concentration was determined for each using the same methodology, as described previously.[23]

The 361 study samples were tested together with 20 serial dilutions (1:2) of a positive control (pool of positive samples at an initial dilution of 1/100) to generate a standard curve for assay quality control, plus 3 technical blanks consisting of Luminex Buffer (1% BSA, 0.05% Tween20, 0.05% sodium azide, 0.005% Triton X-100 in PBS) incubated with beads without samples, to measure non-specific binding of IgG to microspheres. The day of the assay, antigen-coupled microspheres were added in multiplex to the 384-well plate (2000 beads/antigen/well) and mixed with test samples, positive controls and blanks. The loading of microspheres and samples on the plates was performed with a 384-channels Integra Viaflo semi-automatic device and an Integra Assist Plus device with 12-channels Voyager pipette. Final dilution of test samples was 1/250. The plate was incubated for 1 h at room temperature in agitation (Titramax 1000) at 900 rpm and protected from light. Then, the plates were washed three times with 200µL/well of 0.05% Tween 20 in PBS, using a BioTek-405-TS microplate washer (384-well format). Next, 25µL of goat anti-human IgG-phycoerythrin(PE),(GTIG-001,Moss Bio) diluted 1:400 in Luminex buffer were added to each well and incubated for 30 min. The plate was washed again and microspheres resuspended with 80µL of Luminex Buffer, covered with an adhesive film and sonicated 20 seconds on a sonicator bath platform, before acquisition on the Flexmap 3D® reader. At least 50 microspheres per analyte and per well were acquired, and median fluorescence intensity(MFI) was reported as a measure of antigen-specific IgG levels in each serum sample. Raw data was exported with Xponent and managed with R for quality control assessment. Annex 2 shows the standard curves made of 20 serial dilutions of the positive control.

**Annex 3. Standard curve generated with 20 serial dilutions (1:2) of a positive control for quality control**

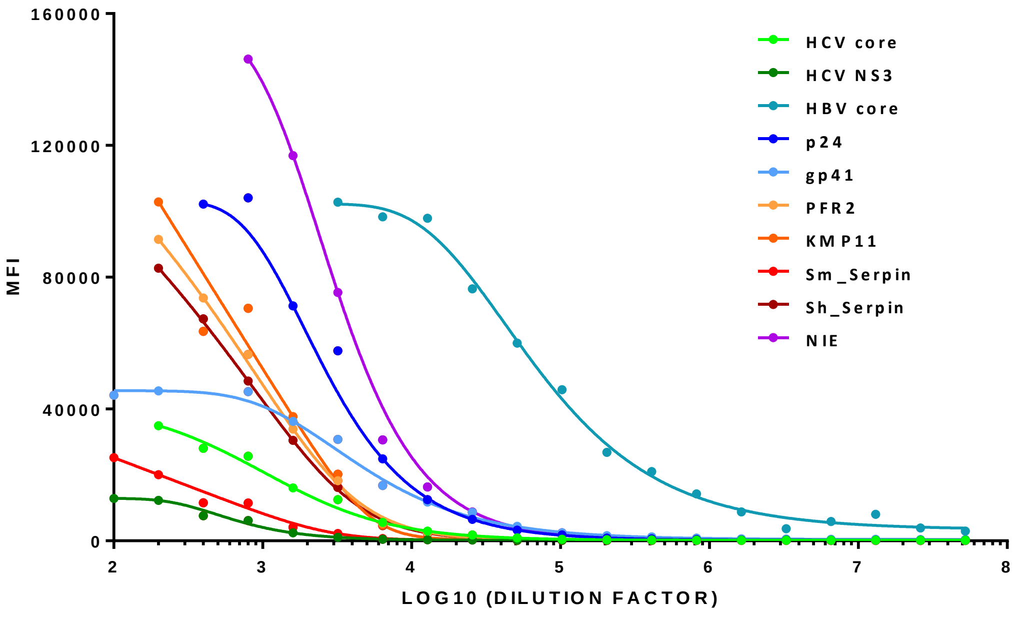


**Annex 4. List of biobanks or collections from which the control samples of the assay have been obtained**

Viral hepatitis: Hospital Clínic Biobank C.0006288

HIV: Samples of the collection registered at the Instituto de Salud Carlos III: C.0000583

*T cruzi*: Samples of the collection registered at the Instituto de Salud Carlos III C.0002610

*Strongyloides*: Sacro Cuore Hospital Tropica Biobank and study STRONG TREAT 1 a 4 (Number Ethical committee Hospital Clínic: 2011-11. EUDRACT number: 2011-002784-24).

*Schistosoma* spp: Sacro Cuore Hospital Tropica Biobank

Healthy controls: Samples of the collection registered at the Instituto de Salud Carlos III: C.0000590

#### Annex 5. STARD checklist

| Section & Topic | No | Item | Reported on page # |
|---|---|---|---|
| **TITLE OR ABSTRACT** | | | |
| | **1** | Identification as a study of diagnostic accuracy using at least one measure of accuracy (such as sensitivity, specificity, predictive values, or AUC) | 1 |
| **ABSTRACT** | | | |
| | **2** | Structured summary of study design, methods, results, and conclusions (for specific guidance, see STARD for Abstracts) | 4 |
| **INTRODUCTION** | | | |
| | **3** | Scientific and clinical background, including the intended use and clinical role of the index test | 6 |
| | **4** | Study objectives and hypotheses | 7 |
| **METHODS** | | | |
| *Study design* | **5** | Whether data collection was planned before the index test and reference standard were performed (prospective study) or after (retrospective study) | 7 |
| *Participants* | **6** | Eligibility criteria | 7-8, 10 |
| | **7** | On what basis potentially eligible participants were identified (such as symptoms, results from previous tests, inclusion in registry) | NA |
| | **8** | Where and when potentially eligible participants were identified (setting, location and dates) | 7-8 |
| | **9** | Whether participants formed a consecutive, random or convenience series | N/A |
| *Test methods* | **10a** | Index test, in sufficient detail to allow replication | 8-9, Annex 2 |
| | **10b** | Reference standard, in sufficient detail to allow replication | 7-8, Annex 1 |
| | **11** | Rationale for choosing the reference standard (if alternatives exist) | 8 |
| | **12a** | Definition of and rationale for test positivity cut-offs or result categories of the index test, distinguishing pre-specified from exploratory | 9 and Annex 3 |
| | **12b** | Definition of and rationale for test positivity cut-offs or result categories of the reference standard, distinguishing pre-specified from exploratory | 8 and Annex 1 |
| | **13a** | Whether clinical information and reference standard results were available to the performers/readers of the index test | 9 |
| | **13b** | Whether clinical information and index test results were available to the assessors of the reference standard | 7 |
| *Analysis* | **14** | Methods for estimating or comparing measures of diagnostic accuracy | 9-10 |
| | **15** | How indeterminate index test or reference standard results were handled | 11 |
| | **16** | How missing data on the index test and reference standard were handled | N/A |
| | **17** | Any analyses of variability in diagnostic accuracy, distinguishing pre-specified from exploratory | 10 |
| | **18** | Intended sample size and how it was determined | NA |
| **RESULTS** | | | |
| *Participants* | **19** | Flow of participants, using a diagram | N/A |
| | **20** | Baseline demographic and clinical characteristics of participants | N/A |
| | **21a** | Distribution of severity of disease in those with the target condition | N/A |
| | **21b** | Distribution of alternative diagnoses in those without the target condition | Annex 8 and Annex 9 |
| | **22** | Time interval and any clinical interventions between index test and reference standard | N/A |
| *Test results* | **23** | Cross tabulation of the index test results (or their distribution) by the results of the reference standard | 10-12 and Table 2 |
| | **24** | Estimates of diagnostic accuracy and their precision (such as 95% confidence intervals) | 10-12 and Table 2 |
| | **25** | Any adverse events from performing the index test or the reference standard | N/A |
| **DISCUSSION** | | | |
| | **26** | Study limitations, including sources of potential bias, statistical uncertainty, and generalisability | 15 |
| | **27** | Implications for practice, including the intended use and clinical role of the index test | 14 |
| **OTHER INFORMATION** | | | |

| | | | |
|---|---|---|---|
| | **28** | Registration number and name of registry | 10 |
| | **29** | Where the full study protocol can be accessed | |
| | **30** | Sources of funding and other support; role of funders | 16 |

#### Annex 6. Seropositivity cutoffs for each of the antigens in the Luminex panel using 2 and 3 SD

| antigen | CUTOFF 2SD | CUTOFF 3SD |
|---|---|---|
| **HCV core** | 3464.5 | 6534.2 |
| **HCV NS3** | 1986.1 | 3181.9 |
| **HBV core** | 35100.5 | 92411.2 |
| **p24** | 10251.2 | 22233.8 |
| **gp41** | 6943.3 | 12035.2 |
| **PFR2** | 38543.2 | 64787.3 |
| **KMP11** | 10159.5 | 23882.9 |
| **Sm_Serpin** | 3541.9 | 7596.9 |
| **Sh_Serpin** | 6460.6 | 15501.0 |
| **NIE** | 14342.5 | 31438.6 |

Cutoff units are MFIs

**Annex 7. Sensitivity and AUC analyses including only confirmed cases with parasitological techniques for each infection as positive controls.**

| | | 2 SD cutoff | | | 3 SD cut-off | | | AUC |
|---|---|---|---|---|---|---|---|---|
| | Antigens | Sensitivity | Specificity | Correctly classified | Sensitivity | Specificity | Correctly classified | |
| Chagas disease[1] | PFR2 | 79.0 | 93.8 | 85.7 | 44.7 | 100 | 70.0 | 0.88 (0.81-0.95) |
| | KMP11 | 97.4 | 96.9 | 97.1 | 89.5 | 100 | 94.3 | 0.99 (0.96-1.00) |
| | Both | 97.4 | 90.6 | 94.3 | 89.5 | 100 | 94.3 | 0.98 (0.95-1.00) |
| *Strongyloides*[2] | NIE | 100 | 90.6 | 94.3 | 100 | 100 | 100 | 1.0 (1.00-1.00) |
| *Schistosoma mansoni*[3] | Sm - serpin | 96.0 | 96.9 | 96.5 | 88.0 | 100 | 94.7 | 0.98 (0.94-1.00) |
| *Schistosoma* haematobium[4] | Sh - serpin | 100 | 96.9 | 97.9 | 93.3 | 96.9 | 95.7 | 0.98 (0.95-1.00) |

1.Excluding indeterminate borderline cases of *T. cruzi* that were confirmed through a third serological test; 2. Including only *Strongyloides* cases detected by stool techniques; 3. Including only confirmed *Schistosoma mansoni* cases; 4. Including only confirmed *Schistosoma haematobium* cases.

**Annex 8. Challenge panel 1: False positives of the Luminex assay in a panel of human samples tested positive for other infections.**

<table>
<tr><th>Negative by Gold standard</th><th>Positive by Gold standard</th><th>False positives (2SD)</th><th>False positives (3SD)</th></tr>
<tr><td rowspan="8">HIV negative control (127)</td><td>T.cruzi (24)</td><td>0</td><td>0</td></tr>
<tr><td>T.cruzi + S.stercoralis (11)</td><td>1</td><td>0</td></tr>
<tr><td>T.cruzi + IgG anti-HBc (4)</td><td>0</td><td>0</td></tr>
<tr><td>S.stercoralis (2)</td><td>1</td><td>0</td></tr>
<tr><td>Schistosoma spp. (18)</td><td>2</td><td>1</td></tr>
<tr><td>Schistosoma spp. + IgG anti-HBc (20)</td><td>5</td><td>2</td></tr>
<tr><td>IgG anti-HBc (24)</td><td>2</td><td>1</td></tr>
<tr><td>IgG anti-HCv (24)</td><td>2</td><td>0</td></tr>
<tr><td rowspan="3">HBV negative control (core) (29)</td><td>T.cruzi (15)</td><td>0</td><td>0</td></tr>
<tr><td>T.cruzi + S.stercoralis (6)</td><td>0</td><td>0</td></tr>
<tr><td>Schistosoma spp. (8)</td><td>0</td><td>0</td></tr>
<tr><td rowspan="9">HCV negative control (81)</td><td>T.cruzi (11)</td><td>0</td><td>0</td></tr>
<tr><td>T.cruzi + S.stercoralis (5)</td><td>0</td><td>0</td></tr>
<tr><td>T.cruzi + IgG anti-HBc (2)</td><td>0</td><td>0</td></tr>
<tr><td>S.stercoralis (2)</td><td>1</td><td>0</td></tr>
<tr><td>S.stercoralis + HIV (1)</td><td>0</td><td>0</td></tr>
<tr><td>Schistosoma spp. (15)</td><td>2</td><td>1</td></tr>
<tr><td>Schistosoma spp. + IgG anti-HBc (11)</td><td>4</td><td>0</td></tr>
<tr><td>IgG anti-HBc (33)</td><td>1</td><td>0</td></tr>
<tr><td>IgG anti-HBc + HIV (1)</td><td>0</td><td>0</td></tr>
<tr><td>T.cruzi negative control (1)</td><td>S.stercoralis (1)</td><td>0</td><td>0</td></tr>
<tr><td rowspan="3">Schistosoma spp. negative control (3)</td><td>T.cruzi (1)</td><td>0</td><td>0</td></tr>
<tr><td>S.stercoralis (1)</td><td>0</td><td>0</td></tr>
<tr><td>S.stercoralis + HIV (1)</td><td>1 (Sm)</td><td>0</td></tr>
<tr><td rowspan="5">S.stercoralis negative control (69)</td><td>T.cruzi (25)</td><td>1</td><td>1</td></tr>
<tr><td>T.cruzi + IgG anti-HBc (4)</td><td>0</td><td>0</td></tr>
<tr><td>Schistosoma spp. (18)</td><td>12</td><td>9</td></tr>
<tr><td>Schistosoma spp. + IgG anti-HBc (21)</td><td>13</td><td>8</td></tr>
<tr><td>IgG anti-HBc + HIV (1)</td><td>1</td><td>1</td></tr>
<tr><td colspan="4">Note: Not all the positive samples stored in biobanks had information on serologies performed for the other infections.</td></tr>
</table>

**Annex 9. Challenge panel 2: False positives of the Luminex assay in a prospective cohort of 48 migrant individuals tested positive for other infections**

| Negative by Gold standard | Positive by Gold standard | False positives (2SD) | False positives (3SD) |
|---|---|---|---|
| HIV negative control (45) | *T.cruzi* (10) | 3 | 1 |
| | *T.cruzi* + *S.stercoralis* (2) | 1 | 0 |
| | *T.cruzi* + IgG anti-HBc (2) | 0 | 0 |
| | *S.stercoralis* (3) | 0 | 0 |
| | *S.stercoralis* + IgG anti-HBc (1) | 0 | 0 |
| | IgG anti-HBc (7) | 0 | 0 |
| | none (20) | 2 | 0 |
| HBV negative control (37) | *T.cruzi* (10) | 0 | 0 |
| | *T.cruzi* + *S.stercoralis* (2) | 0 | 0 |
| | *S.stercoralis* (3) | 0 | 0 |
| | HIV* (1) | 1 | 1 |
| | none (21) | 3 | 1 |
| HCV negative control (48) | T.cruzi (10) | 2 | 1 |
| | *T.cruzi* + *S.stercoralis* (2) | 0 | 0 |
| | *T.cruzi* + IgG anti-HBc (2) | 0 | 0 |
| | *S.stercoralis* (3) | 0 | 0 |
| | *S.stercoralis* + IgG anti-HBc (1) | 0 | 0 |
| | IgG anti-HBc (8) | 0 | 0 |
| | HIV* (1) | 0 | 0 |
| | none (21) | 0 | 0 |
| *T.cruzi* negative control (16) | *S.stercoralis* (3) | 0 | 0 |
| | none (13) | 0 | 0 |
| *Schistosoma* spp. negative control (22) | *T.cruzi* (5) | 1 (Sh) | 0 |
| | *T.cruzi* + *S.stercoralis* (2) | 0 | 0 |
| | *T.cruzi* + IgG anti-HBc (1) | 0 | 0 |
| | *S.stercoralis* + IgG anti-HBc (1) | 0 | 0 |
| | IgG anti-HBc (7) | 1 (Sm) & 1 (Sh) | 1 (Sm) & 1 (Sh) |
| | none (6) | 1 (Sm) & 2 (Sh) | 0 |
| *S.stercoralis* negative control (38) | *T.cruzi* (10) | 2 | 0 |
| | *T.cruzi* + IgG anti-HBc (2) | 0 | 0 |
| | IgG anti-HBc (7) | 2 | 1 |
| | HIV* (1) | 0 | 0 |
| | none (18) | 1 | 0 |

*HIV gold standard serology result was indeterminate; no positive cases for HCV and *Schisotosoma* spp. Infection were reported in the cohort.
